# Enhancing Patient Safety in Opioid Prescribing: A Systematic Review of Potential Indicators

**DOI:** 10.1101/2023.12.07.23299686

**Authors:** Neetu Bansal, Wael Y. Khawagi, Nan Shang, Li-Chia Chen

## Abstract

**Background:** This systematic review aimed to identify a comprehensive list of prescribing safety indicators for opioids in any setting from published literature.

**Methods:** Literature that reported prescribing indicators from 1990-2019 was retrieved from a published systematic review. A subsequent search was conducted from seven electronic databases to identify additional studies from 2019 to 2022. Potential opioid safety prescribing indicators were extracted from studies that reported prescribing indicators of non-injectable opioids and narcotics prescribed to adults with or without specific conditions, concomitant medications, or laboratory monitoring with concerns about the potential safety risk of harm. Retrieved indicators were split by each opioid and merged for the same drugs. Identified indicators were categorised by the type of problem, medication, patient condition/disease, and the risk of the indicators.

**Results:** Of the 107 indicators retrieved from 48 included articles, 71 were included. Thirty-five (49.3%) opioid prescribing indicators focused on a specific class of opioids, i.e., ‘opioids’ (n=30, 42.3%) and ‘strong opioids’ (n=5, 7.0%); tramadol and fentanyl were the most commonly reported drug (n=6, 8.5%). The indicators account for six types of problems: medication inappropriate to the population (n=16), omission (n=7), inappropriate duration (n=4), inadequate monitoring (n=7), drug-disease interaction (n=16), and drug-drug interaction (n=27). Of all indicators, older age (over 65) is the most common risk factor (n=34, 47.9%). Central nervous system-related adverse effects are the risk of concern for the 27 indicators associated with drug-drug interaction (n=24, 88.9%). Besides, five of the six ‘omission’ indicators are related to ‘without using laxatives’.

**Conclusion:** This review identified a comprehensive list of indicators that can be applied to flag patients with a high risk of opioid-related harm to facilitate complex decision-making in optimising opioids for pain management. Further research is needed to validate and determine the feasibility of identifying hazardous prescribing in various care settings.

## Introduction

Opioids, one type of potent analgesic, remain the mainstay approach for treating moderate to severe pain used acutely after surgery and for cancer-related pain (1, 2). However, in recent decades, opioid analgesics have been increasingly used in patients with chronic non-cancer pain in Western countries (3). Chronic pain, commonly referred to as pain lasting for three or more months affects between one-third and one-half of the United Kingdom (UK) population (4, 5). Due to the complex mechanisms of pain physiology and pathology, multimodal biopsychosocial treatments, including non-pharmacological options, are recommended for managing chronic primary pain rather than analgesics alone (6). Yet the marked increase in opioids used for chronic pain has become a public health concern (7).

There is a lack of evidence to support the long-term effectiveness of opioids for managing chronic pain (8); instead, much literature has revealed dose-dependent adverse effects and the risk of severe harm, including the potential of drug abuse and addiction associated with long-term opioid use (8, 9). Opioid safety is of particular concern in some vulnerable patient groups (10), such as older people, because age-related pharmacokinetic and pharmacodynamic changes may increase the sensitivity to adverse drug effects (11). Besides, older patients with chronic pain may also have multiple co-morbidities and become potential polypharmacy candidates, consequently having a higher risk of adverse drug-drug interactions (11, 12). Therefore, it is judicious to have a system to identify and review patients on long-term opioids to manage chronic pain.

Prescribing quality indicators are widely used in identifying the effectiveness of prescribing problematic or inappropriate polypharmacy as part of the quality indicators of healthcare services by allowing relevant stakeholders, e.g., health boards, primary care clusters, general practices and prescribers, to compare their current prescribing practice against an agreed quality standard (13, 14). Furthermore, prescribing safety indicators focusing on potentially hazardous prescribing and inadequate medication monitoring practices that place patients at risk of harm can be used to monitor prescribing safety and prevent prescribing-related harm (14). The development of prescribing indicators should be evidence-based, transparent, easily understood, and ideally, validated by a group of experts using consensus methodology (15).

Although numerous sets of prescribing quality and safety indicators and inappropriate prescribing criteria have been developed for different populations and settings there is no consensus on opioid safety prescribing indicators to prevent potentially hazardous consequences. Therefore, to develop evidence-based opioid safety prescribing indicators, this systematic review aimed to identify explicit indicators or criteria in the existing literature related to opioid prescribing that could potentially be used to assess prescribing safety in adults.

## Materials and methods

This review followed the principles of Preferred Reporting Items for Systematic Reviews and Meta-analyses (PRISMA) guidance (16). The protocol for this systematic review was registered with PROSPERO (CRD42022343776).

### Eligibility criteria

This review included studies that reported developing, validating or updating a set of explicit indicators or criteria that assess prescribing safety. After that, studies that reported prescribing indicators of non-injectable opioids and narcotics prescribed to adults (18 years and over) with or without specific conditions, concomitant medications, or laboratory monitoring with concerns about the potential risk of harm were included. The prescribing safety indicators should explicitly include terms related to opioids prescribed to the target population or drug-drug or drug-disease oriented terms with concerns about the risk of harm (17). Three drug product databases, the American Society of Health-System Pharmacists Drug Information (18), British National Formulary (19) and Martindale (20), were referred to in order to define opioid and narcotics medications.

Studies were excluded if they exclusively focused on children and adolescents (under 18 years), patients with cancer-related pain or receiving palliative care, or injectable opioids. Besides, studies that exclusively reported the incidence of existing prescribing indicators in the clinical setting or reported older versions for included indicators were excluded. Furthermore, studies that reported implicit indicators (i.e., not drug-specific) only or quality indicators not explicitly focused on the risk of harm (e.g., patients older than 65 years already on long-acting opioids with breakthrough pain and not on short-acting opioids) were also excluded.

### Information sources and literature search strategy

Firstly, literature that reported explicit prescribing indicators from 1990-2019 was retrieved from a systematic review published by Khawagi *et al*. (2019) (13), and then a subsequent update review was conducted to identify studies that reported prescribing indicators from 2019 to 30 September 2022.

Khawagi *et al.*’s search strategy aimed to identify explicit prescribing indicators of any kind. They used a combination of three sets of search terms: medication safety terms, quality measure terms and indicators development/validation terms. The authors retrieved 79 articles that reported any prescribing indicators published from 1990 to 2019 with no restrictions on the publication language, type of study design, focused country, targeted setting, or targeted population (13). Khawagi *et al.’s* review is a good data source for identifying the studies from 1990-2019 since the Beers criteria, one of the earliest criteria to describe inappropriate prescribing was published in 1991 (21).

An update of the systematic review was conducted by applying the same search strategy and search terms used by Khawagi *et al*. (13) on the electronic databases, including Medline, Embase, PsycINFO, Web of Science, Health Management Information Consortium (HMIC), International Pharmaceutical Abstracts (IPA), and Cumulative Index to Nursing and Allied Health Literature (CINAHL) to identify articles published from 2019 to 30 September 2022 (Appendix 1). The identified records were imported into the reference management programme (EndNote 20, Calrivate), where the duplications were moved. The remaining records were screened and selected in the following processes.

### Study selection

Titles and abstracts of the literature identified from the electronic database search were screened independently by two reviewers (WK, NS) to identify literature for developing, validating, or evaluating any prescribing indicators or criteria. If there were discrepancies in the inclusion decision of a study, then it was progressed to full-text screening. Two reviewers (WK, NS) further reviewed the full texts of eligible articles independently according to inclusion and exclusion criteria and reviewers’ clinical knowledge to identify the eligible studies that reported any prescribing indicators. Any discrepancy was resolved by discussing with a third reviewer (LCC or NB) to reach a consensus. The reasons for exclusion were documented. Finally, the full-text articles identified from Khawagi *et al.’s* review (n=79) and the updated systematic review were scrutinised to include studies that reported opioid-specific prescribing safety indicators.

### Data extraction

The data of included studies were extracted by two reviewers (NS, WK) using an electronic data collection form and double-checked by two reviewers (NB, LCC). Three major categories of data were extracted from the literature, including (1) literature information: title, leading author, country, and year; (2) methodology: study design, setting, targeted population, indicators sources, and validation methods; and (3) opioid-related prescribe indicators: total number of indicators, opioids name of the medication, duration of medication, disease condition, concomitant medication, risk of the prescribing.

### Data synthesis and analysis

Several principles were imposed to transform each indicator extracted from the included literature into a statement in the following standard format.

*“Prescribing opioids [class or medicine] +/- with other drugs [class or medicine] +/- to a patient +/- aged [age in years] +/- with disease [disease condition] (+/- for the risk of conditions [potential opioid-related harm]).”*

Afterwards, duplicate indicators were removed. In addition, if an indicator included more than one opioid, it was split into multiple indicators. For example, “Prescribing propoxyphene or pentazocine to patients over 65 years” was split into two indicators.

In addition, the indicators were categorised into six prescribing problem categories. (Medication inappropriate to population, drug-disease interaction, drug-drug interaction, inappropriate duration, inadequate monitoring, and omission) (Table 1). These categories were adapted from previous studies (13, 22–24). A summarised compact version of the included indicators was also provided, combining similar overlapped indicators. A descriptive analysis of the above items was selected, and numbers and portions were calculated when appropriate.

**Table 1.** Definitions of the six types of prescribing problems.

| Type of indicator | Prescribing problem |
| --- | --- |
| **Medication inappropriate to the population** | Medication that is potentially prescribed inappropriately to a specific population. |
| **Omission** | Medication should be prescribed with a specific diagnosis, condition, or prescription. |
| **Inappropriate duration** | Medication that was prescribed with an inappropriate duration. |
| **Inadequate monitoring** | Medications that were not monitored adequately. |
| **Drug-disease interaction** | Medication that is potentially prescribed inappropriately with a specific diagnosis or condition. |
| **Drug-drug interaction** | Medication that potentially interacts with another medication. |

(Note) Seventy-one indicators were identified. The complete list can be found in Appendix 2.

## Results

### Selection of studies

Of the 9,387 records identified from the electronic database search, 3,353 duplicate and 921 ineligible records (older than 2019 were removed) (Figure 1). After screening the titles and abstracts of 5,113 records, 5,047 were excluded for not including prescribing indicators and the remaining 66 records were considered for full-text review. Overall, 32 articles published from 2019 to 2022 that reported any prescribing indicators were selected after excluding 34 articles. The reasons for exclusion included studies that did not develop prescribing indicators (n=20), studies that applied published indicators (n=7), adapted, or translated one tool to another country (n=2), reported duplicate data (n=1) or were not published in full text (i.e., conference abstract; n=4).

**Figure 1.**
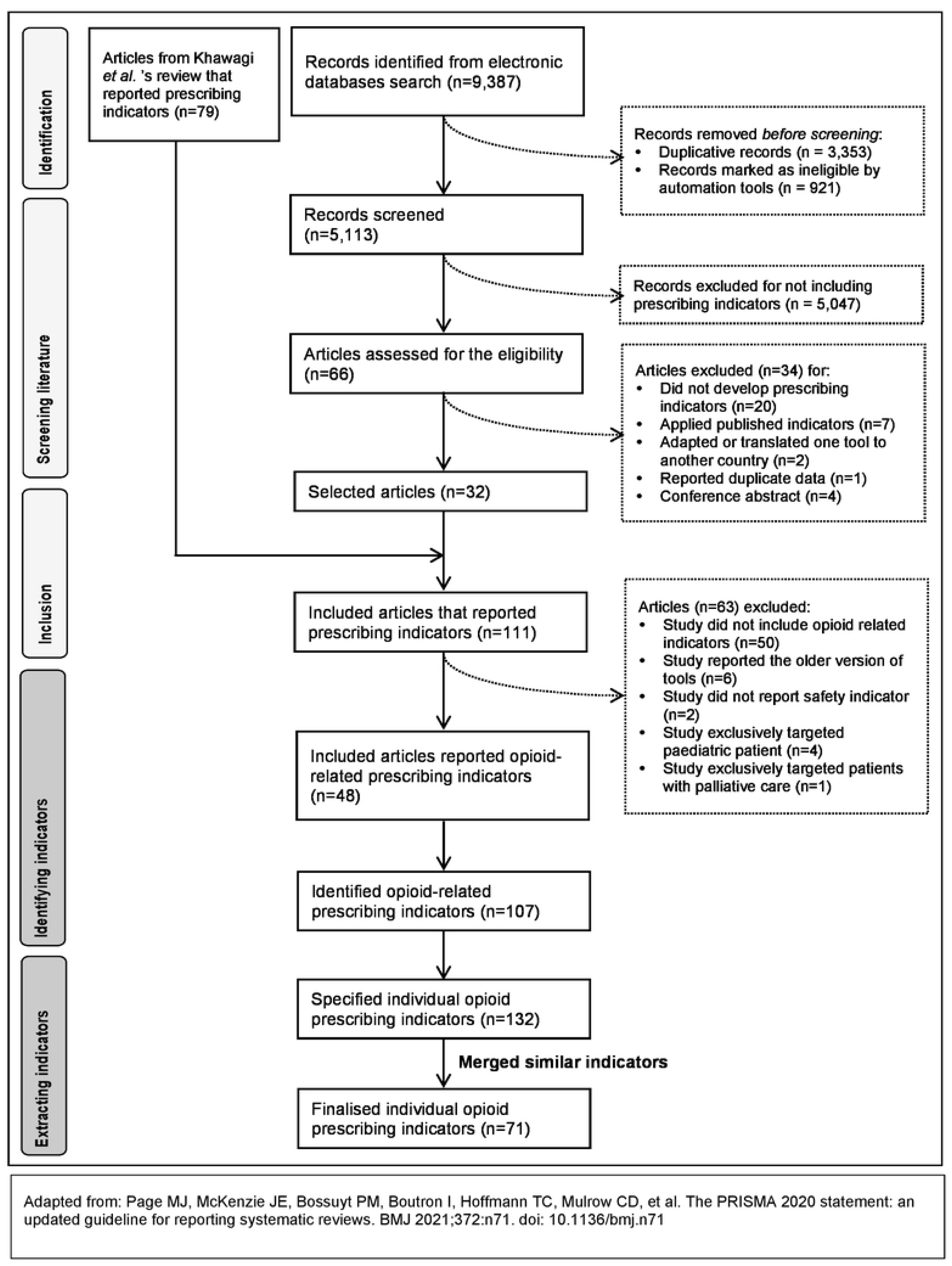
PRISMA flow diagram on process of selecting literature and retrieving opioid prescribing indicators.

In total, 111 articles were included after combining the 32 articles with 79 studies from Khawagi *et al.*’s review (13). The full texts of these 111 articles reported any prescribing indicators were reviewed, and 48 articles that reported opioid-related prescribing indicators were included in this systematic review. The 63 articles were excluded as these studies did not include opioid-related indicators (n=50), reported the older version of indicators (n=6), did not report any safety indicators (n=2), or exclusively focused on paediatric (n=4) or patients with palliative care (n=1).

### Characteristics of the included articles

#### Publication year and countries

The 48 included studies which reported at least one opioid-related prescribing indicator (2, 9, 24–69) were published during the period 2010-2019 (n=24)(13, 27, 28, 30–33, 35, 37–39, 41, 43, 45, 46, 49, 50, 54, 55, 57, 60, 63, 65, 66) followed by 2000-2009 (n=13) (9, 24, 29, 34, 42, 44, 48, 56, 58, 59, 62, 67, 68) and 2020-2022 (n=10) (2, 25, 26, 36, 40, 51–53, 61, 69) and only one published in 1990-1999 (47) (Table 2).. Furthermore, except for one study that developed indicators for international use (31), most studies (n=23) aimed to develop indicators to be used in Europe (2, 9, 25, 26, 30, 33, 35–40, 43, 46, 48, 50, 53, 54, 57–59, 62, 65) followed by North America (n=12) (24, 27, 32, 42, 44, 47, 52, 55, 56, 64, 66, 68), Asia (n=7) (29, 41, 51, 60, 61, 67, 69), Australia (n=3) (28, 34, 63) and South America (n=2)(45, 49).

**Table 2.**
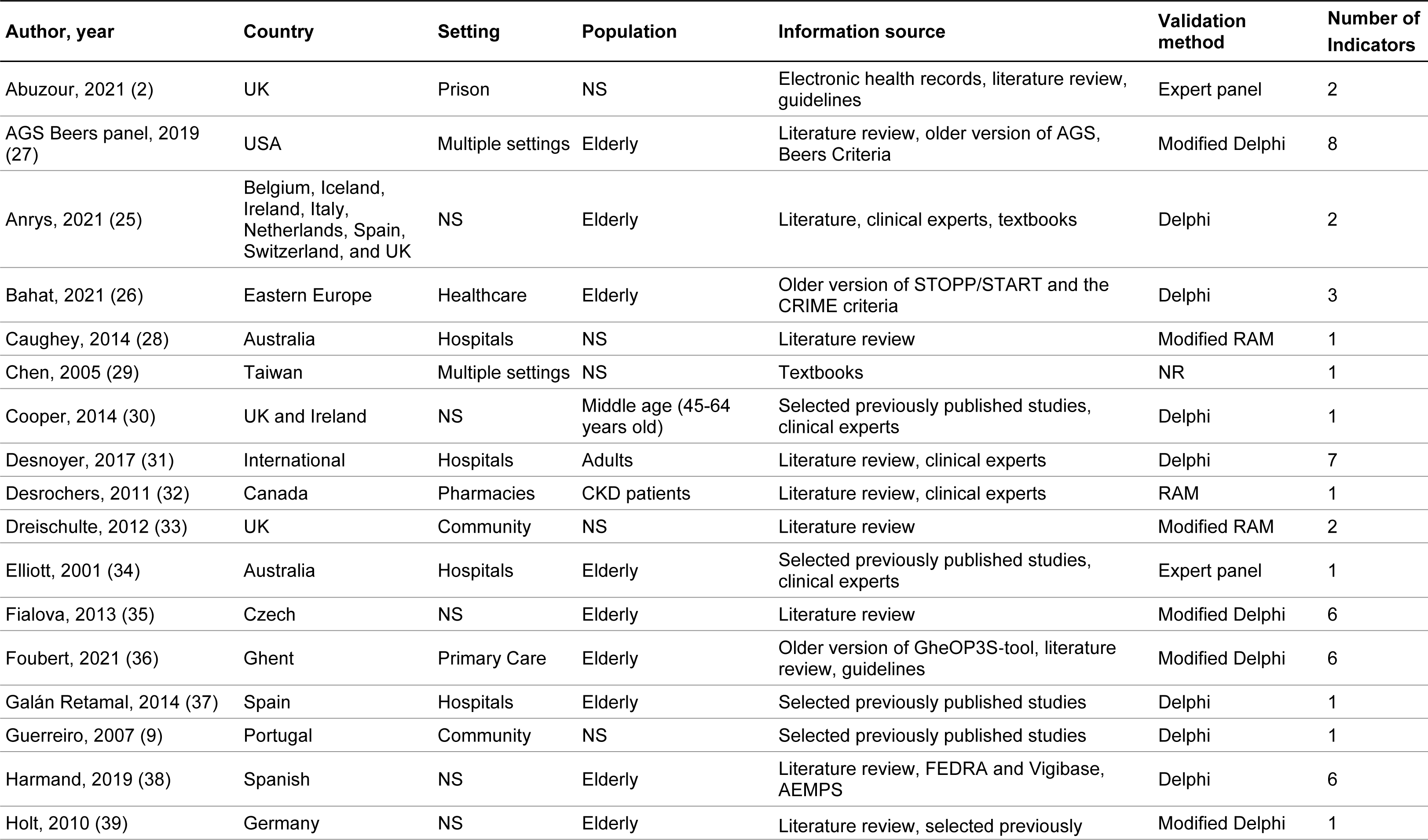
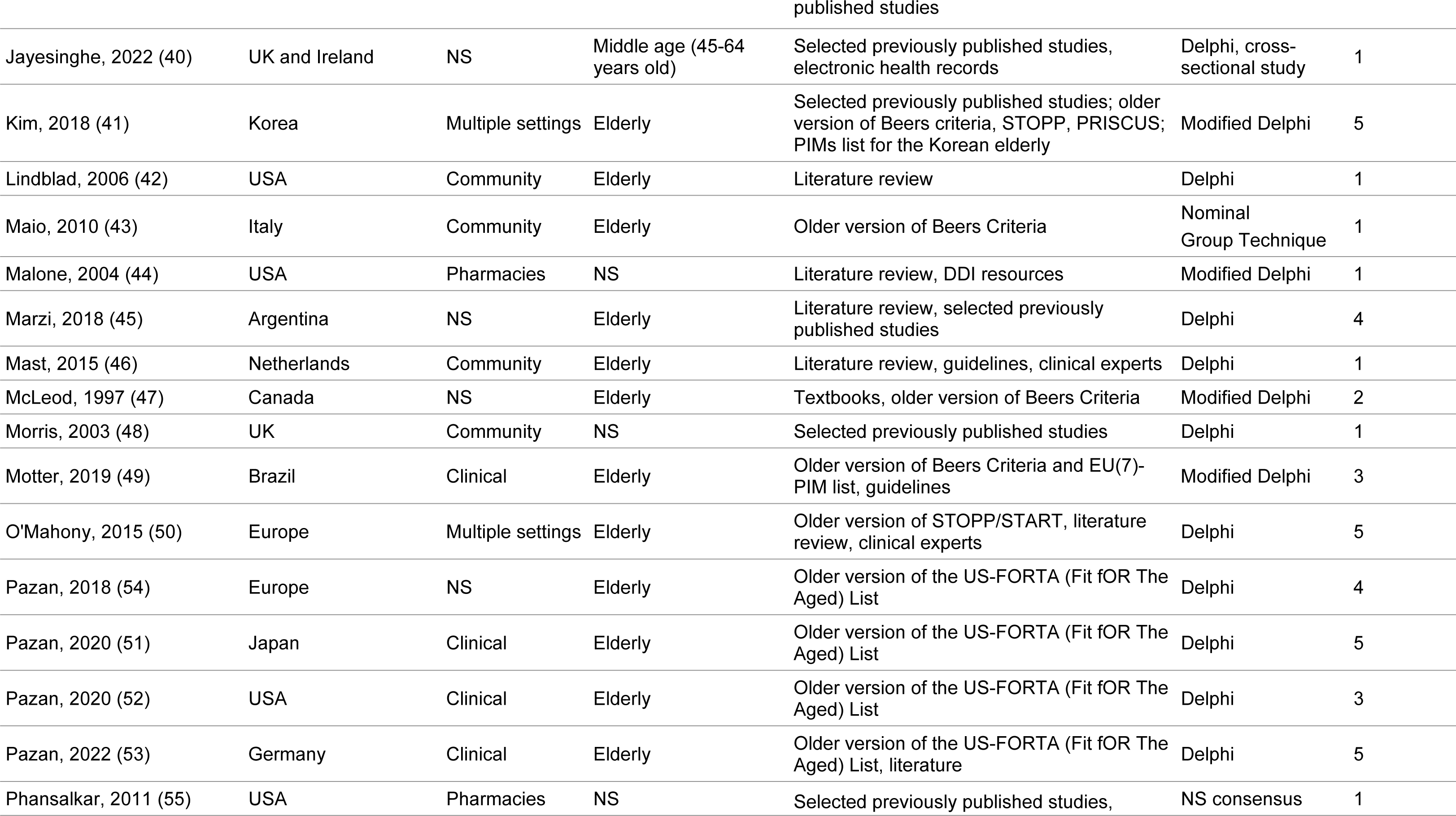
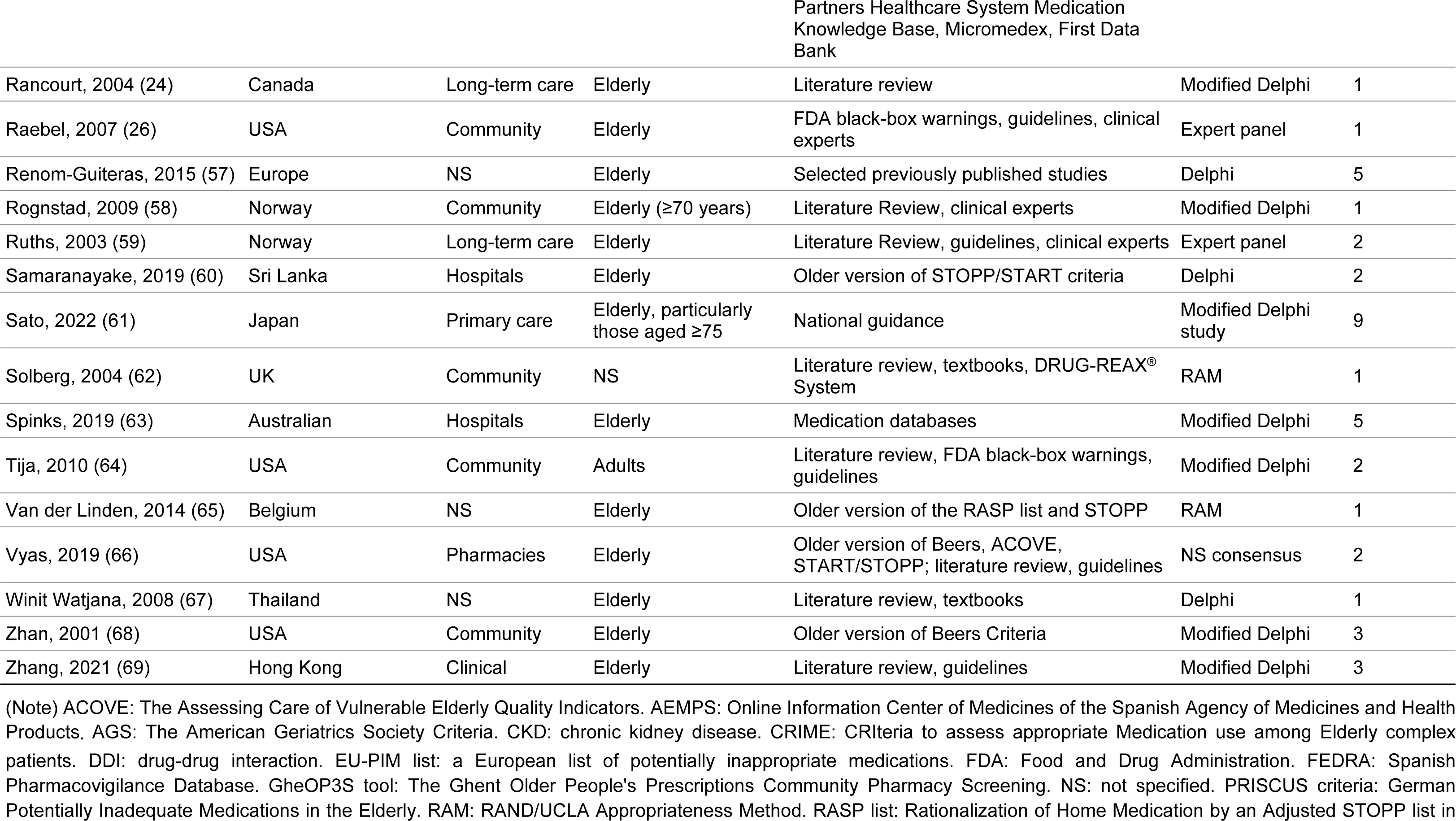
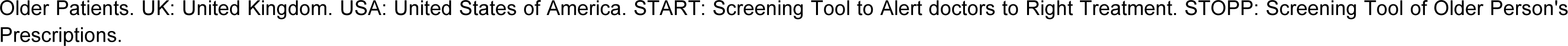
Summary of included studies.

#### Target population and setting

Although nine (21%) of the 48 studies did not specify the target population(2, 9, 28, 29, 33, 44, 48, 55, 62), older people were the most common target population (n=34; 71%). The elderly population was primarily defined as age ≥65 years old (n=32) (24–27, 34–39, 41–43, 45–47, 49–54, 56, 57, 59, 60, 63, 65–69) except for one study that defined elderly as age ≥70 (57) and another as ≥75 years old (58). In addition, although adults (n=2) (31, 64), middle-aged patients (n=2) (30, 40) or patients with chronic kidney disease (n=1) (32) were also investigated, these studies did not specify the age in defining the study population, except for two studies defined the middle age as 45-64 years old (30, 40).

Of the 48 studies, 18 (38%) did not specify the setting to apply the indicators (25, 26, 30, 35, 38–40, 45, 47, 49, 51–54, 57, 65, 67, 69) and four (8.3%) implemented the indicators in multiple settings (27, 29, 41, 50). The remaining 26 studies focused on applying the indicators in various settings, including hospitals (n=6) (28, 31, 34, 37, 60, 63), primary care (general practices) (n=2, 4.2%) (36, 61), community pharmacy (n=4) (32, 44, 55, 66), long-term care setting (care home or nursing home) (n=2) (24, 59), prison (n=1, 2.9%) (2), and any patients in the community (n=11, 22.9%) (9, 33, 42, 43, 46, 48, 56, 58, 62, 64, 68).

#### Method to identify and validate prescribing indicators

Most of the studies (n=29, 60.4%) retrieved the indicators from multiple sources or strategies (2, 25, 27, 30–32, 34, 36, 38–41, 44–50, 53, 55, 56, 58, 59, 62, 64, 66, 67, 69) and 19 (39.6%) studies only adapted one information source or strategy (9, 24, 26, 28, 29, 33, 35, 37, 42, 43, 51, 52, 54, 57, 60, 61, 63, 65, 68). Literature review (n=25, 52.1%)(2, 24, 25, 27, 28, 31–33, 35, 36, 38, 39, 42, 44–46, 50, 53, 58, 59, 64, 66, 67, 69, 70) or referring to an old version of indicators (n=17, 35.4%) (26, 27, 36, 41, 43, 47–54, 60, 65, 68, 70) were the most used strategies to retrieve opioid-related indicators.

Except for one (2.9%) study (29), all studies reported the validation method of prescribing indicators, and the most used is the Delphi method (n=34, 70.8%). The remaining studies used the RAND/UCLA appropriateness method (RAM) (n=5, 10.4%) (28, 32, 33, 65, 70) expert panel method (n=4, 8.3%) (2, 34, 56, 59), non-specific consensus method (n=2, 4.2%) (55, 66) combined Delphi and cross-sectional study (n=1, 2.1%) (40) and nominal group technique (n=1, 2.1%) (43).

In addition, various types of consensus methodologies were applied. For example, of the 34 studies that adopted a Delphi consensus approach, 20 studies reported a Delphi method (9, 25, 26, 30, 31, 37, 38, 40, 42, 45, 46, 48, 50–54, 57, 60, 67) and 15 applied a modified Delphi method (24, 27, 35, 36, 39, 41, 44, 47, 49, 58, 61, 63, 64, 68, 69), Similarly, of the five studies that reported using the RAM method, two applied a modified RAM (2/5=40.0%) (28, 32, 33, 62, 65).

### Categories of opioid-related prescribing indicators

Overall, 107 original opioid prescribing indicators were identified from the 48 studies. Of these, 14 indicators (13.1%) that involved more than one opioid medicine were split by each opioid, resulting in 132 indicators. After merging indicators for the same drugs or items of the same drug class, 71 opioid-related prescribing indicators were identified. Of the 71 indicators, 35 (49%) focused on the classification of opioids using the terms ‘opioids’ (n=30, 42%) and ‘strong opioids’ (n=5, 7%). In the remaining 36 indicators, 16 opioid medications, tramadol, fentanyl, and pethidine, were the most reported drugs, each for six indicators (8.5%) (Table 3).

**Table 3.** Opioid analgesics and categories reported across six types of prescribing problems.

| Opioid drug | A | B | C | D | E | F | Subtotal |
| --- | --- | --- | --- | --- | --- | --- | --- |
| Opioids | 1 | 5 |  | 1 | 7 | 16 | 30 |
| Strong opioids | 1 | 2 | 2 |  |  |  | 5 |
| Buprenorphine | 1 |  |  |  |  |  | 1 |
| Codeine |  |  | 1 |  |  |  | 1 |
| Dextromethorphan/quinidine | 1 |  |  |  |  |  | 1 |
| Dextropropoxyphene | 1 |  |  |  |  | 1 | 2 |
| Fentanyl | 1 |  |  |  | 2 | 3 | 6 |
| Methadone | 1 |  |  |  |  |  | 1 |
| Morphine | 1 |  |  |  | 2 |  | 3 |
| Narcotic antitussives | 1 |  |  |  |  |  | 1 |
| Oxycodone/naloxone | 1 |  |  |  |  |  | 1 |
| Oxycodone |  |  |  |  |  | 1 | 1 |
| Pentazocine | 1 |  | 1 |  |  |  | 2 |
| Pethidine | 1 |  |  |  | 3 | 2 | 6 |
| Propoxyphene | 1 |  |  |  |  | 1 | 2 |
| Tapentadol | 1 |  |  |  |  |  | 1 |
| Tilidine/naloxone | 1 |  |  |  |  |  | 1 |
| Tramadol | 1 |  |  |  | 2 | 3 | 6 |
| **Subtotal** | 16 | 7 | 4 | 1 | 16 | 27 | 71 |

(Note) A: medication inappropriate to the population, B: omission, C: inappropriate duration, D: inadequate monitoring, E: drug-disease interaction, F: drug-drug interaction.

When classifying the 71 indicators into six types of prescribing problems, drug-drug interaction was the most common type (n=27; 38.0%), followed by drug-disease interaction (n=16; 22.5%), prescribing to an inappropriate population (n=16; 22.5%), omission (n=7; 9.9%), inadequate monitoring (n=7; 9.9%) and inappropriate duration (n=4; 5.6%). Within each group, similar indicators were further grouped to provide a simplified summary of the pivotal characteristics of the indicators (Table 4).

**Table 4.**
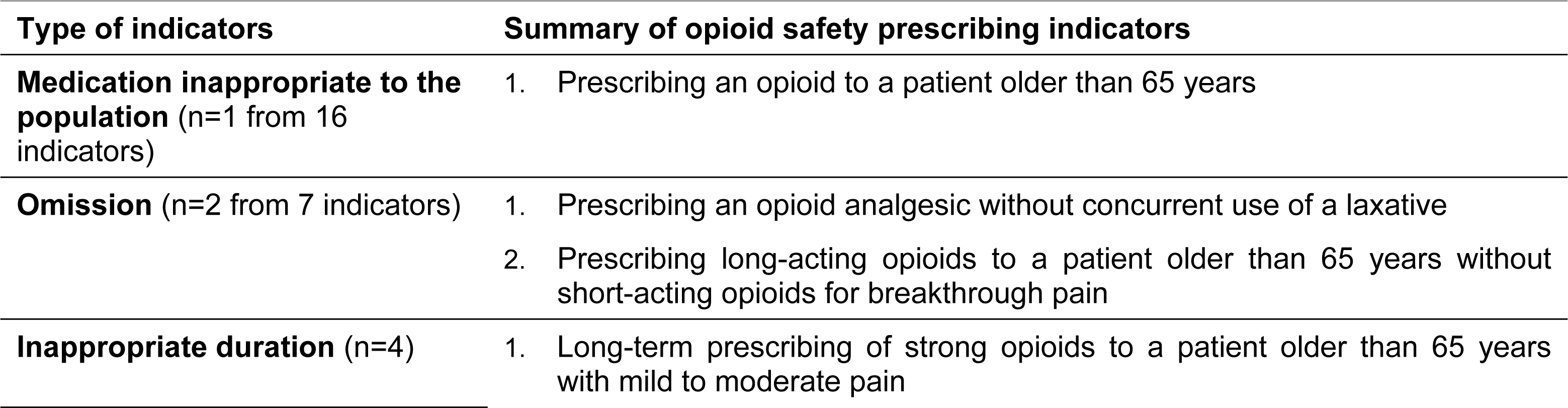
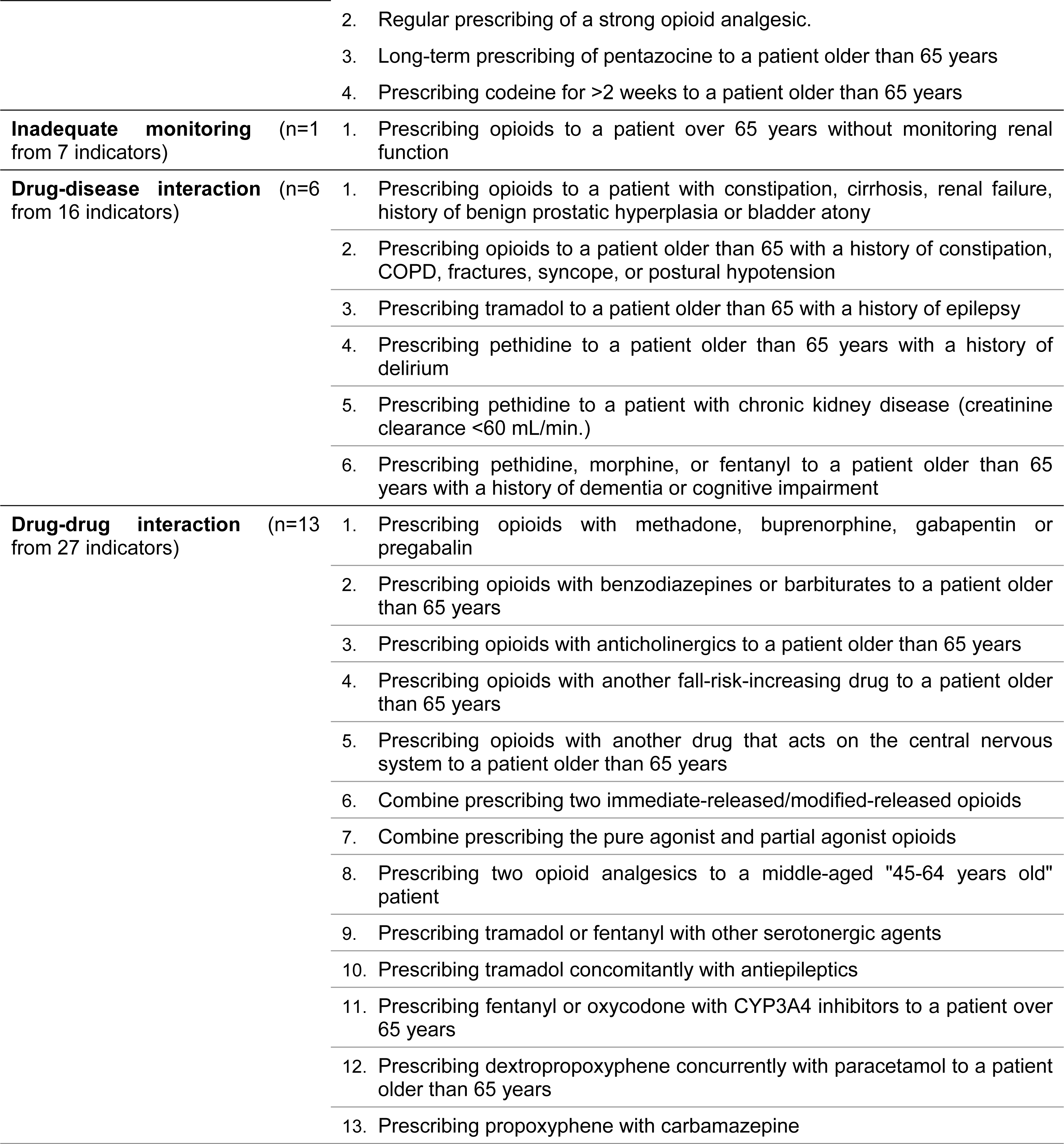
Summary of opioid safety prescribing indicators.

| Type of indicators | Summary of opioid safety prescribing indicators |
| --- | --- |
| **Medication inappropriate to the population** (n=1 from 16 indicators) | 1. Prescribing an opioid to a patient older than 65 years |
| **Omission** (n=2 from 7 indicators) | 1. Prescribing an opioid analgesic without concurrent use of a laxative<br>2. Prescribing long-acting opioids to a patient older than 65 years without short-acting opioids for breakthrough pain |
| **Inappropriate duration** (n=4) | 1. Long-term prescribing of strong opioids to a patient older than 65 years with mild to moderate pain |

|  |  |  |
| --- | --- | --- |
|  |  | 2. Regular prescribing of a strong opioid analgesic.<br>3. Long-term prescribing of pentazocine to a patient older than 65 years<br>4. Prescribing codeine for >2 weeks to a patient older than 65 years |
| **Inadequate monitoring** (n=1 from 7 indicators) |  | 1. Prescribing opioids to a patient over 65 years without monitoring renal function |
| **Drug-disease interaction** (n=6 from 16 indicators) |  | 1. Prescribing opioids to a patient with constipation, cirrhosis, renal failure, history of benign prostatic hyperplasia or bladder atony<br>2. Prescribing opioids to a patient older than 65 with a history of constipation, COPD, fractures, syncope, or postural hypotension<br>3. Prescribing tramadol to a patient older than 65 with a history of epilepsy<br>4. Prescribing pethidine to a patient older than 65 years with a history of delirium<br>5. Prescribing pethidine to a patient with chronic kidney disease (creatinine clearance <60 mL/min.)<br>6. Prescribing pethidine, morphine, or fentanyl to a patient older than 65 years with a history of dementia or cognitive impairment |
| **Drug-drug interaction** (n=13 from 27 indicators) |  | 1. Prescribing opioids with methadone, buprenorphine, gabapentin or pregabalin<br>2. Prescribing opioids with benzodiazepines or barbiturates to a patient older than 65 years<br>3. Prescribing opioids with anticholinergics to a patient older than 65 years<br>4. Prescribing opioids with another fall-risk-increasing drug to a patient older than 65 years<br>5. Prescribing opioids with another drug that acts on the central nervous system to a patient older than 65 years<br>6. Combine prescribing two immediate-released/modified-released opioids<br>7. Combine prescribing the pure agonist and partial agonist opioids<br>8. Prescribing two opioid analgesics to a middle-aged "45-64 years old" patient<br>9. Prescribing tramadol or fentanyl with other serotonergic agents<br>10. Prescribing tramadol concomitantly with antiepileptics<br>11. Prescribing fentanyl or oxycodone with CYP3A4 inhibitors to a patient over 65 years<br>12. Prescribing dextropropoxyphene concurrently with paracetamol to a patient older than 65 years<br>13. Prescribing propoxyphene with carbamazepine |

#### Drug-drug interaction

The 27 drug-drug interaction indicators identified in this review can be summarised into 16 groups (Table 4). These indicators described opioids as a class or specific drug interacting with another therapeutic class (e.g., prescribing opioids with a *tricyclic antidepressant* to patients over 65 years), with medications that pose the same risk (e.g., prescribing opioids with Page 18 of 30 another *fall-risk-increasing drug* to patients aged >65 years), with specific medications (e.g., prescribing an opioid with *gabapentin* or *pregabalin*) or with another opioid (e.g., combine prescribing of *pure agonist* and *partial agonist* opioids). Most drug-drug interactions are due to the concern of pharmacological effects on the central nervous system.

#### Drug-disease interaction

The 16 indicators focused on drug-disease interactions were summarised in six groups (Table 4). These referred to opioids prescribed to patients with varying conditions, such as current constipation, history of constipation, or benign prostatic hyperplasia (Figure 2). Besides, four of the six indicators’ groups specified prescribing for patients over 65 (Table 4).

**Figure 2.**
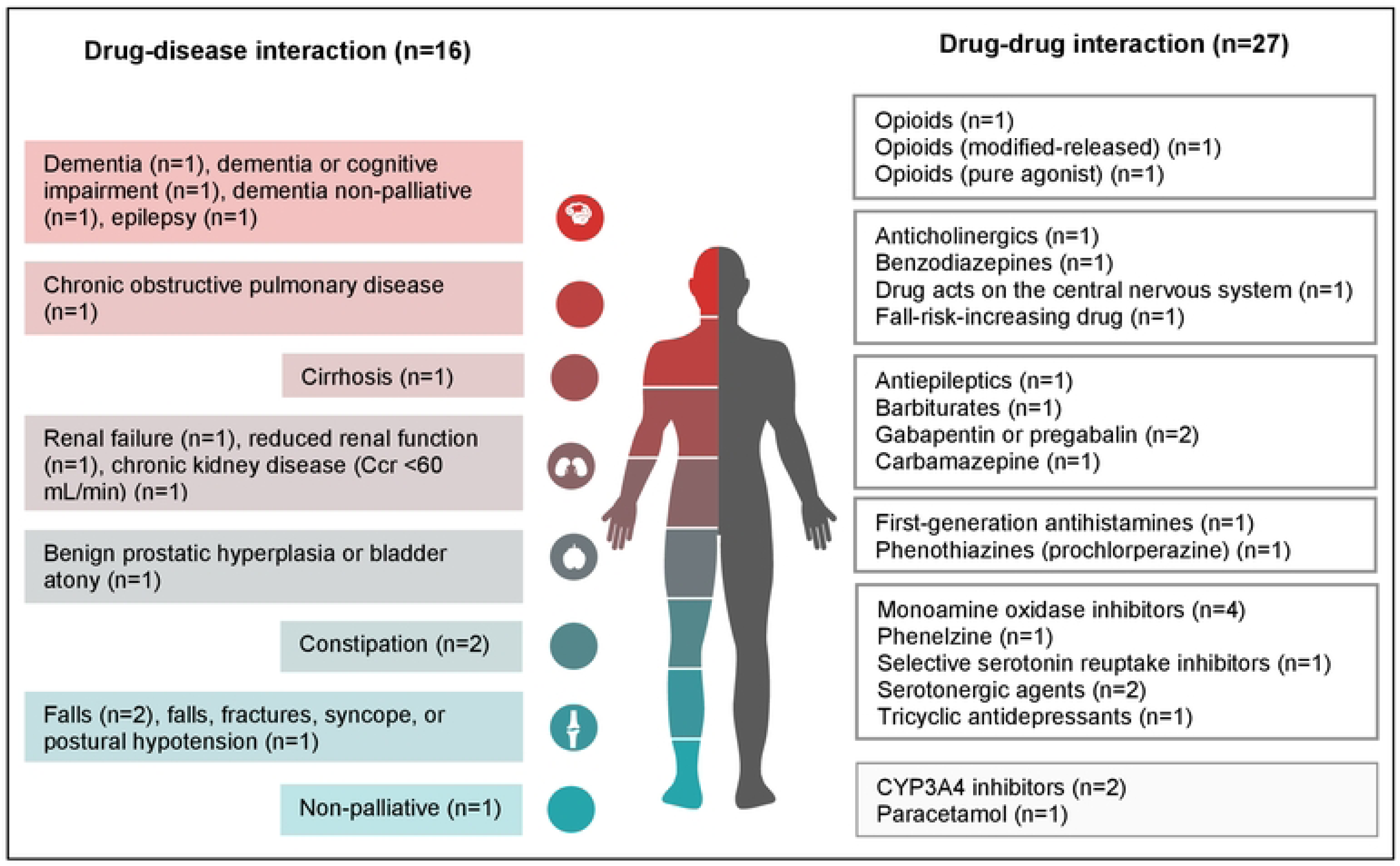
Indicators concerning the prescription problems of drug-disease or drug­ drug interactions.

#### Prescribing to inappropriate population

All the 16 indicators of “prescribing to an inappropriate population” were related to prescribing opioids to patients older than 65 years (Table 4). Of which, only one indicator referred to opioids being prescribed as *a class* for patients over 65 years. The remaining 14 indicators referred to a specific opioid prescribed to the same elderly population (Appendix 2).

#### Omission

Six of the seven omission indicators described patients being prescribed opioids as a class without being prescribed a laxative. The remaining indicator described prescribing long-acting opioids to patients older than 65 years without short-acting opioids for breakthrough pain (Table 4). Of the six indicators related to omitting a laxative, four specified that the prescribing was for patients over the age of 65 years, two specified prescribing the opioid over two weeks, more than four or long-term, and two specified prescribing strong opioids (Appendix 2).

#### Inappropriate duration

Of the four indicators focused on inappropriate duration, two described patients being prescribed strong opioids as a class, concerning the long-term prescribing of strong opioids to patients over 65 years with mild to moderate pain or regular prescribing of strong opioid analgesics (Table 4). The remaining two indicators referred to prescribing a specific opioid, including prescribing codeine for more than two weeks to patients over 65 years or long-term prescribing of pentazocine to patients older than 65 years (Table 4).

#### Inadequate monitoring

Only one indicator described Inadequate monitoring. This indicator described prescribing opioids to patients over 65 years without monitoring their renal function.

## Discussion

This study reported published opioid prescribing indicators that could potentially be used to assess prescribing safety for adults. These indicators can be further validated to assess hazardous prescribing across clinical settings. Most of the indicators identified are based on expert consensus and clinical guidelines, reflecting the implicit knowledge and best practices that healthcare professionals have developed. One notable advantage of these indicators is their ability to flag all potential hazardous risks. However, it is crucial to consider their limitations. Some indicators may lack robust supporting evidence, such as the prevalence of adverse consequences linked to specific scenarios. Consequently, this may lead to challenges in implementation, for example, persuading practitioners or patients to change their practices. Furthermore, quantifying the risk of poor outcomes remains difficult, presenting challenges in measuring the effectiveness of implementing these indicators (71). The risk associated with opioids concerning these indicators is not remarkably different from general considerations for other medications. However, the risk and severity of the consequences may vary when comparing opioids with other drugs. It is prudent to note that advanced age alone is not necessarily a cause of concern. Nevertheless, older adults may pose an increased risk due to the pharmacodynamic changes associated with ageing, pre-existing co-morbidities and polypharmacy, which can be an issue in this age group (10). Specific patient populations, such as those with chronic pain, warrant special attention, as the prevalence of chronic pain is notably higher amongst older adults, alongside other co-existing medical conditions(72). In the case of patients managing chronic pain, they often find themselves contending with other symptoms, leading to the potential for polypharmacy (73).

Among the identified prescribing problems, drug-drug interactions were identified as the most common prescribing problem, emphasising the need for vigilance in identifying and managing potential interactions when prescribing opioids. Within this category, various opioids (either as a class or specific drugs) should be highlighted. This finding aligns with previous studies highlighting the risk of adverse events associated with polypharmacy, particularly in the older population, due to multimorbidity, polypharmacy, and age-related changes in pharmacokinetics and pharmacodynamics (74). Prescribing indicators targeting drug-drug interactions can serve as valuable tools to guide healthcare professionals in optimising medication regimes and minimising harm (70).

Most concerns related to polypharmacy primarily revolve around central nervous system (CNS) implications, emphasising the need to prioritise vigilance regarding severe consequences such as CNS depression (75). This finding reflects the well-known potential of opioids to cause central nervous system-related adverse effects such as sedation, respiratory depression, and cognitive impairment (76) Nevertheless, the complexity of pain conditions calls for individualised risk management and optimisation of decision-making, rooted in a patient-centred approach rather than solely relying on pre-defined indicators (77). Similarly, drug-disease interactions raise concerns regarding the potential risks for patients with a history of certain diseases. Interpreting the risk and formulating appropriate responses poses a challenge, necessitating a consensus approach to applying these indicators appropriately (78).

In addition to class-specific indicators, many indicators considered the patient’s accompanying conditions, diseases, or medical history. The most prevalent condition-related indicator was “opioids prescribed without laxatives”, highlighting the significance of considering potential adverse effects with detrimental impacts on patients’ quality of life and medication safety when prescribing opioids to patients with specific conditions. Compared to drug-drug, drug-disease interaction, and inappropriate population, the indicators for the omission, inappropriate duration and monitoring are more specific and seem more straightforward to implement (as the mitigation response to those scenarios). Nevertheless, the most common omission, whether to prescribe a laxative, should still be based on patients’ conditions, so a patient-centred care approach and shared decision-making should be considered (79). Shared decision-making relies on patients and clinicians using the best available evidence (79). This finding also suggests the importance of a patient-centred approach to prescribing, accounting for individual patient characteristics and medical histories to optimise opioid safety.

The development and use of prescribing quality and safety indicators are crucial in improving healthcare quality and preventing prescribing-related harm (80). However, the absence of consensus on opioid safety prescribing indicators for primary care settings highlights the need for evidence-based indicators to guide prescribing practices and enhance patient safety. The findings of this review provide a valuable starting point for developing evidence-based opioid safety prescribing indicators that can be further validated and implemented in clinical practice.

Our comprehensive approach in including studies with published prescribing indicators that cover all opioids and the most updated list of opioid-related indicators is a strength of this study. Our rigorous review process, which involves researchers specialised in methodology (WK and LCC) and a specialist practitioner (NB) reviewing identified indicators, adds confidence to the validity and reliability of our results. However, we acknowledge this review has some limitations. The search was limited to published English literature, which may introduce publication bias and exclude relevant unpublished studies or grey literature. Additionally, the inclusion criteria focused on implicit indicators and excluded studies that reported indicators not focused on the risk of harm. Consequently, there is a possibility that some potentially relevant indicators may have been overlooked. Nevertheless, this review provides a comprehensive overview of the current literature on prescribing indicators for opioid and serves as a valuable resource.

## Conclusion

This systematic review identified a range of potential opioid safety prescribing indicators from the published literature. These indicators can contribute to developing evidence-based prescribing practices and preventing potential harm associated with opioid use. Further validation using relevant healthcare data and implementation of these indicators in different clinical settings are necessary to enhance patient safety and optimise opioid prescribing practices. Future research should also explore the feasibility and effectiveness of these indicators and consider their integration into clinical guidelines and decision-support systems to support healthcare professionals in delivering safe and effective opioid therapy.

## Data Availability

All relevant data are within the manuscript and its Supporting Information files.

## Authors’ contribution

LCC conceptualised, oversaw, planned, and managed this project. WK and NB developed the methodology and registered the protocol. NS and WK conducted the search, screening, data extraction, management, and analysis. All authors are involved in data interpretation and writing this manuscript.

